# Assessment of Knowledge and Attitudes Toward HPV, Cervical Cancer, and Vaccination Barriers among Undergraduates in Ghana: A Cross-Sectional Study

**DOI:** 10.64898/2026.06.25.26356537

**Authors:** Isaac Aidoo Erzuah, Babah Abdulrahman, Elwin Kordai Quarshie, Anna Eresong Doosogla, Corban Enam Bubutor, Mimijoy Aba Erzuah, Amidu Alhassan, Christiana Asiedu

## Abstract

**Background:** Human papillomavirus (HPV) infection is a leading cause of cervical cancer globally, disproportionately affecting women in developing countries like Ghana. Despite the recent introduction of national HPV vaccination programs, vaccine uptake among young adults remains suboptimal. This study aimed to assess the levels of knowledge, attitudes, and perceived barriers toward HPV, cervical cancer, and vaccination among undergraduate students in Ghana to inform future public health interventions.

**Methods:** A cross-sectional study was conducted among 699 undergraduate students at the University of Cape Coast, Ghana. A multistage stratified random sampling technique was employed to ensure disciplinary representation. Data were collected using a validated, semi-structured digital questionnaire covering socio-demographics, knowledge of HPV, attitudes toward vaccination, and perceived barriers. Descriptive statistics were utilized to summarize findings. Chi-square tests were performed to assess bivariate associations, and binary logistic regression analysis was conducted to identify predictors of good knowledge and positive attitudes toward vaccination, with statistical significance set at p < 0.05.

**Results:** 51.9% of students demonstrated good knowledge of HPV and vaccination. A significant gender disparity was observed: while male students displayed higher levels of clinical knowledge, female students held significantly more positive attitudes toward vaccination (p < 0.05). Major barriers included profound social stigma, with 77.9% of students expressing concern over partner perception and 65.6% reporting embarrassment regarding the association between the vaccine and sexually transmitted infections. Misconceptions were prevalent, with 46.6% of participants incorrectly believing the vaccine could cure existing infections.

**Conclusion:** A clear knowledge-attitude gap exists among Ghanaian undergraduates, complicated by pervasive psychosocial barriers. Current vaccine delivery models, which often center on reproductive health or STI clinics, inadvertently reinforce stigma. To improve vaccination coverage, public health initiatives must transition toward a stigma-neutral model of care that integrates HPV immunization into routine primary health services, framing it as a preventive cancer-fighting strategy rather than a sexual health intervention.

## Introduction

Human papillomavirus (HPV) infection is the most prevalent sexually transmitted infection (STI) globally, with clinical manifestations ranging from benign cutaneous lesions to life-threatening malignancies(1). Persistent infection with high-risk HPV genotypes is the primary etiological factor for cervical cancer, a significant global public health crisis (1,2). While cervical cancer is the fourth most common malignancy among women worldwide, it disproportionately burdens developing nations; in Ghana, it ranks as the second most common cancer and a leading cause of cancer-related mortality (2,3). Current surveillance indicates approximately 3,000 new cases and 1,800 deaths annually in Ghana (4), necessitating urgent, evidence-based preventive action.

The HPV vaccine is a cornerstone of primary cervical cancer prevention. By targeting high-risk genotypes, specifically HPV 16 and 18, which are responsible for the majority of global cases, vaccination offers a highly efficacious strategy for disease reduction (5,6). A study examining the distribution of high-risk HPV genotypes among cervical cancer cases in Ghana, reported common genotypes including HPV types 59, 35, 18, 16, and 33 (7). These findings highlight the importance of vaccination as a primary preventive strategy to reduce the burden of HPV-related diseases.

Ghana launched its first nationwide human papillomavirus (HPV) vaccination campaign in October 2025 through the Ghana Health Services, in partnership with the Vaccine Alliance (Gavi), the World Health Organization (WHO), and United Nations Children’s Fund (UNICEF) (4). This initiative aligns with the WHO global strategy to eliminate cervical cancer as a public health problem by 2030, which targets 90% of girls being fully vaccinated against HPV by age 15, 70% of women being screened by ages 35 and 45, and 90% of women with cervical disease receiving appropriate treatment(1). The introduction of HPV vaccination in Ghana therefore represents an important step toward translating global commitments into national action in the fight against cervical cancer. As of 2025, there are 8 licensed HPV vaccines, five of which have received WHO pre-qualification and are available globally(1). Ghana has introduced Gardasil vaccine into hospitals, schools and communities, where it is available nationwide (8). These vaccines have demonstrated high efficacy in preventing HPV infections and related cervical lesions when administered prior to exposure to the virus, typically recommended for preadolescents and young adults(4).

Despite ongoing efforts by the Ministry of Health to expand HPV vaccination coverage across Ghana, several barriers continue to limit optimal uptake. These include limited awareness and knowledge about cervical cancer and screening practices (9–12), as well as vaccine hesitancy, which significantly influences vaccination intentions and acceptance (13–15).

University students constitute a high-priority demographic for these interventions. As young adults approaching or within their peak sexually active years, they remain an important target group for vaccination before potential exposure to HPV(16). However, the effectiveness of HPV vaccination programs depends not only on vaccine availability but also on adequate knowledge, positive attitudes and perceived barriers (8). Previous studies have identified knowledge about HPV infection and HPV vaccination as important predictors of vaccine uptake(8,17).

Currently, there is a lack of comprehensive data on how Ghanaian university students perceive the intersection of HPV awareness and vaccination. Addressing this knowledge-attitude gap is essential for refining public health strategies. Therefore, this study aimed to assess the knowledge, attitudes, and perceived barriers regarding HPV and cervical cancer vaccination among Ghanaian undergraduates to provide an evidence base for targeted interventions.

## Methods

### Study Design and Setting

This cross-sectional study was conducted among undergraduate students at the University of Cape Coast, Ghana. The University was chosen for the study because it has culturally diverse groups of students, both local and international, within their reproductive ages expected to have different opinions regarding the subject under study. The university has a hospital and a large, diverse student population, providing an appropriate setting for assessing HPV-related knowledge, attitudes and vaccination willingness among young adults. The University has approximately 45,000 undergraduate students. It has several colleges, including the College of Humanities and Legal Studies, College of Education Studies, College of Agriculture and Natural Sciences, and the College of Health and Allied Sciences. These colleges host students from diverse academic backgrounds, providing an appropriate setting for examining knowledge, attitudes, and acceptance of HPV vaccination among young adults. The study population consisted of undergraduate students enrolled across different academic levels (Level 100–600). University students represent a critical population for HPV prevention research, as many fall within the recommended age range for HPV vaccination, education and can play an important role in disseminating health information within their communities.

### Eligibility Criteria

The study included undergraduate students at the University, who are aged 16 years or older. Participants were eligible if they were available during data collection, willing to participate and provided informed consent to participate in the study.

### Sample Size Determination

The sample size was calculated using the Cochran’s sample size formula (18):

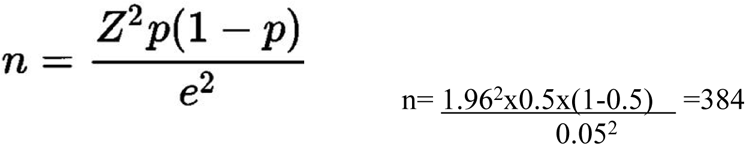

Where:

n = required sample size

Z = z-score corresponding to 95% confidence level (1.96)

p = estimated response distribution (0.5 is used to maximize sample size)

e = margin of error (0.05 or 5%)

Using these assumptions, the minimum sample size required was 384 participants. To improve statistical power and account for potential non-response or incomplete questionnaires, the sample size was increased. A total of 699 students ultimately participated in the study.

### Sampling Technique

A multistage stratified random sampling technique was employed. First, colleges were selected via simple random sampling. Second, faculties and departments were randomly selected within each college to ensure disciplinary representation. Third, one academic level (100–600) was randomly selected per department. Finally, students were recruited using systematic random sampling based on departmental rosters, with a sampling interval calculated to ensure proportional representation across departments.

### Data Collection Tools and Procedure

Data was collected using a semi-structured questionnaire designed to assess knowledge, attitudes, and barriers to HPV, cervical cancer and HPV vaccination. Data was collected using digital formats such as; Google Forms. Data collection was conducted from 25^th^ May 2026 to 18^th^ June 2026. Participation was strictly voluntary and informed consent was obtained digitally; participants indicated their consent by selecting the’I Agree’, which was a mandatory prerequisite for accessing the survey instrument.

A questionnaire was designed consisting of four (4) sections: Section A determined participant socio-demographic information. Section B assessed awareness of HPV infection, HPV vaccine and cervical cancer. Section C determined knowledge of HPV infection and HPV vaccine. Section D assessed attitudes towards HPV vaccination. The attitude questions included positive and negative statements about HPV vaccination. Negatively worded items were reverse coded to ensure that higher scores consistently indicated more positive attitudes. The questionnaire was adapted from existing studies(19,20). Participants were informed about the purpose of the study and assured of the confidentiality and anonymity of their responses. Students who agreed to participate completed the questionnaire voluntarily. No identifying information was collected, ensuring anonymity and minimizing social desirability bias.

### Pretesting

A pilot study was conducted with 40 students to assess item clarity and feasibility. Data from the pilot were excluded from the final analysis. Cronbach’s alpha coefficients for the knowledge and attitude sections were 0.771 and 0.845, respectively, indicating acceptable internal consistency.

### Data Analysis and Measurement of Variables

Data collected was entered into Microsoft Excel spreadsheet and analyzed using IBM SPSS Statistics version 27.0. Descriptive statistics including frequency distributions were used to summarize participants’ sociodemographic characteristics, knowledge, attitudes, and vaccination-related behaviors. Factors associated with knowledge and attitude were assessed using the Chi-square (χ²) test. Binary regression analysis was conducted to identify predictors of good HPV knowledge and positive attitude. Regression results were reported as regression coefficients (B) for linear models and odds ratios (ORs) with 95% confidence intervals (CIs) for logistic models. Model adequacy for logistic regression was verified using the events-per-variable (EPV) criterion, with an EPV greater than 12 considered sufficient for reliable estimates(21). All statistical tests were set at a significance p <.05.

Knowledge of HPV infection, cervical cancer, and HPV vaccination was assessed using 21 true/false items. Each correct response was scored as “1”, while an incorrect response was scored as 0, producing a total knowledge score ranging from 0 to 21, with higher scores indicating better knowledge. Regarding knowledge scores, a cutoff point of 75% (answering 16 of the 21 questions) was represented good knowledge about HPV infection and vaccine(22,23). Attitudes toward HPV vaccination were assessed using 17 items rated from 1 (strongly disagree) to 5 (strongly agree). Negatively worded items were reverse coded to ensure that higher scores consistently indicated more positive attitudes. The observed scores ranged from 25 to 63. Attitude was categorized into negative and positive attitude. A score ≥ median was positive attitude (22,24).

## Results

### Sociodemographic and Behavioral Characteristics

The sociodemographic and behavioral characteristics of the study population (N = 699) are summarized in **Table 1**. The minimum and maximum ages of respondents were 16 and 36 years respectively with a mean ± SD of 21 ± 2.58 years. Majority of the participants were single 87.1% (n=609). Slightly more than half of the respondents were males 54.1% (n=378). Participants were drawn from different academic colleges within the University, including the College of Health and Allied Sciences 26.8% (n=187), Humanities and Legal Studies 25.5%(n=178), Education Studies 24.2%(n=169), and Agriculture and Natural Sciences 23.6%(n=165). Students across all academic levels were represented, with the highest proportion from Level 600 19.2% (n=134).

**Table 1:**
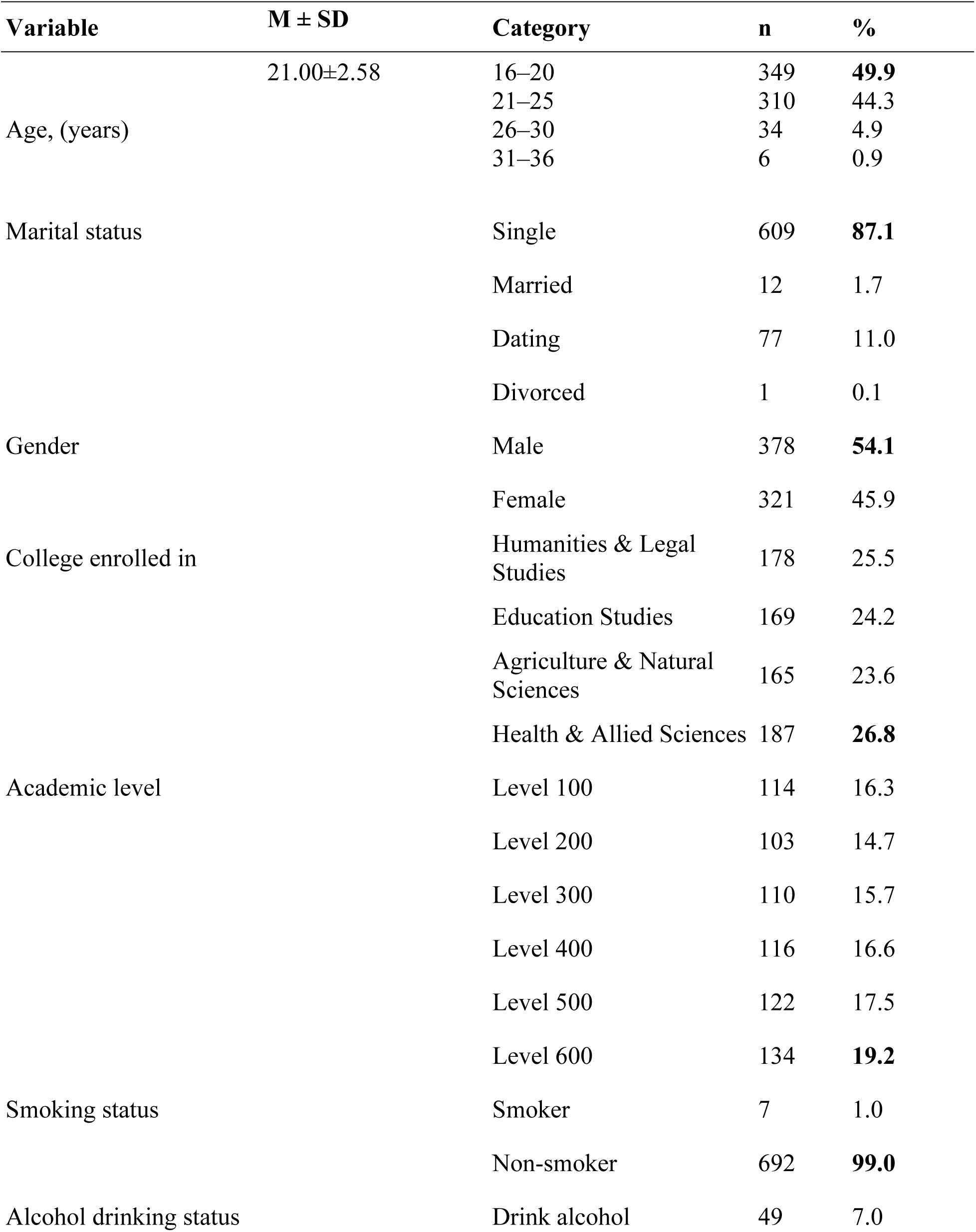
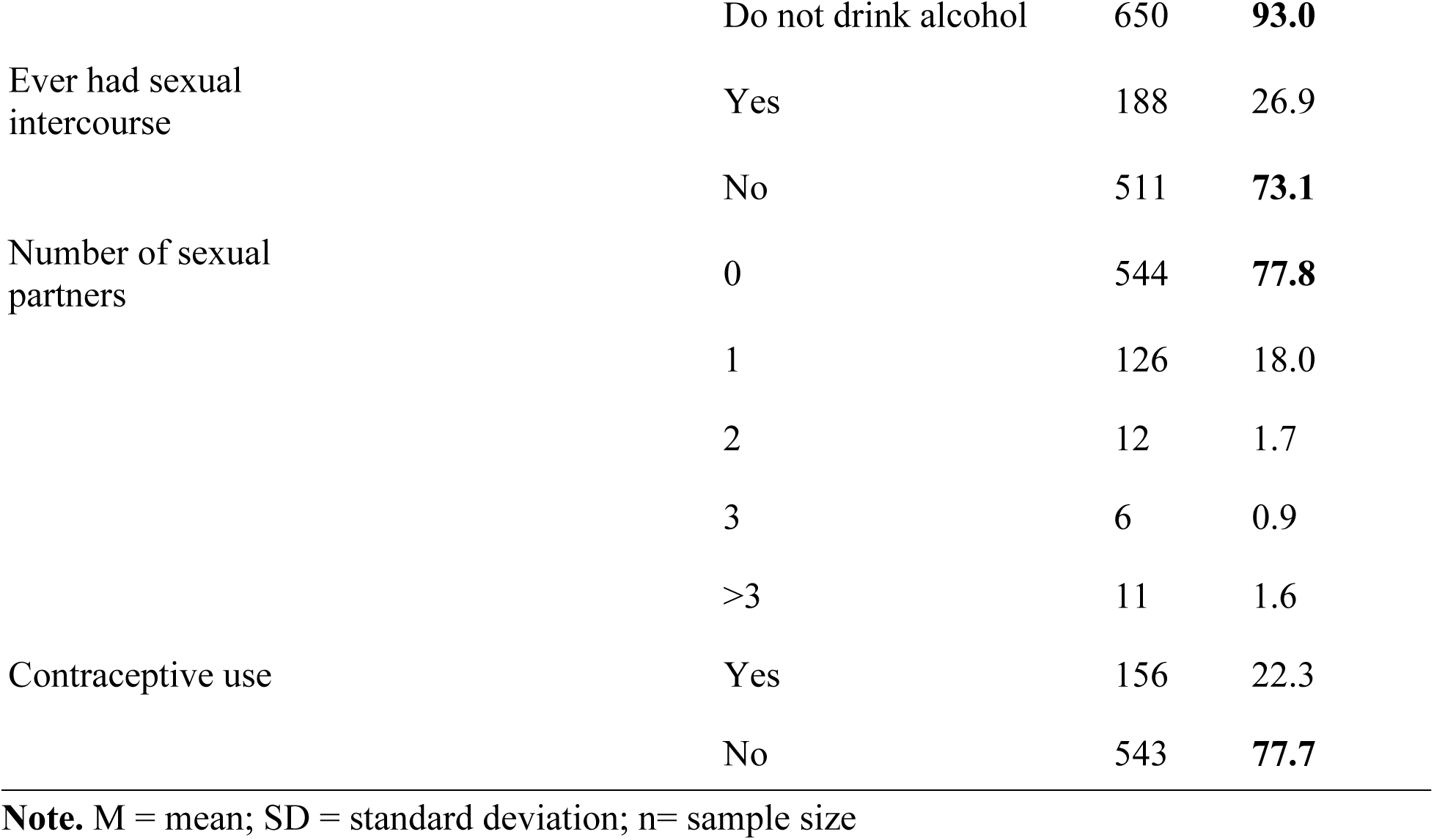
Sociodemographic Characteristics of Respondents (N = 699)

| **Variable** | **M ± SD** | **Category** | **n** | **%** |
| --- | --- | --- | --- | --- |
| Age, (years) | 21.00±2.58 | 16–20 | 349 | **49.9** |
|  |  | 21–25 | 310 | 44.3 |
|  |  | 26–30 | 34 | 4.9 |
|  |  | 31–36 | 6 | 0.9 |
| Marital status |  | Single | 609 | **87.1** |
|  |  | Married | 12 | 1.7 |
|  |  | Dating | 77 | 11.0 |
|  |  | Divorced | 1 | 0.1 |
| Gender |  | Male | 378 | **54.1** |
|  |  | Female | 321 | 45.9 |
| College enrolled in |  | Humanities & Legal Studies | 178 | 25.5 |
|  |  | Education Studies | 169 | 24.2 |
|  |  | Agriculture & Natural Sciences | 165 | 23.6 |
|  |  | Health & Allied Sciences | 187 | **26.8** |
| Academic level |  | Level 100 | 114 | 16.3 |
|  |  | Level 200 | 103 | 14.7 |
|  |  | Level 300 | 110 | 15.7 |
|  |  | Level 400 | 116 | 16.6 |
|  |  | Level 500 | 122 | 17.5 |
|  |  | Level 600 | 134 | **19.2** |
| Smoking status |  | Smoker | 7 | 1.0 |
|  |  | Non-smoker | 692 | **99.0** |
| Alcohol drinking status |  | Drink alcohol | 49 | 7.0 |

|  |  |  |  |
| --- | --- | --- | --- |
|  | Do not drink alcohol | 650 | **93.0** |
| Ever had sexual intercourse | Yes | 188 | 26.9 |
|  | No | 511 | **73.1** |
| Number of sexual partners | 0 | 544 | **77.8** |
|  | 1 | 126 | 18.0 |
|  | 2 | 12 | 1.7 |
|  | 3 | 6 | 0.9 |
|  | >3 | 11 | 1.6 |
| Contraceptive use | Yes | 156 | 22.3 |
|  | No | 543 | **77.7** |

**Note.** M = mean; SD = standard deviation; n= sample size

Regarding behavioral characteristics, majority of participants were non-smokers, 99.0% (n=692), and 93.0% (n=650) reported that they did not consume alcohol. 73.1% (n=511) reported no sexual experience. In addition, 77.8% (n=544) reported having no sexual partners, and 77.7%(n=543) indicated that they did not use contraceptives (**Table 1)**

### Knowledge of HPV, Cervical Cancer and HPV Vaccination

Participants’ knowledge regarding HPV, cervical cancer, and vaccination is detailed in **Table 2**. While general awareness was high with 93.8% (n=656) having heard of cervical cancer, 84.0% (n=587) of genital warts, and 77.5% (n=542) of HPV, there was a notable deficit in policy-level awareness, as 59.5% (n=416) of participants were unaware of current WHO recommendations on HPV vaccination.

**Table 2:** Knowledge of HPV, Cervical Cancer, and HPV Vaccination (N = 699)

| **Item** | **Response** | **n** | **%** |
| --- | --- | --- | --- |
| Heard of HPV | Yes | 542 | **77.5** |
|  | No | 157 | 22.5 |
| Heard of cervical cancer | Yes | 656 | **93.8** |
|  | No | 43 | 6.2 |
| Heard of genital warts | Yes | 587 | **84.0** |
|  | No | 112 | 16.0 |
| Heard of HPV vaccine | Yes | 508 | **72.7** |
|  | No | 191 | 27.3 |
| Aware of WHO recommendations | Yes | 283 | 40.5 |
|  | No | 416 | **59.5** |
| Cervical cancer is associated with HPV | True | 609 | **87.1** |
|  | False | 90 | 12.9 |
| HPV is an STI | True | 619 | **88.6** |
|  | False | 80 | 11.4 |
| >50% of people infected in lifetime | True | 488 | **69.8** |
|  | False | 211 | 30.2 |
| Transmitted only when symptoms present | True | 215 | 30.8 |
|  | False | 484 | **69.2** |
| HPV causes oropharyngeal cancer | True | 517 | **74.0** |
|  | False | 182 | 26.0 |
| Specific HPV types cause cervical cancer | True | 567 | **81.1** |
|  | False | 132 | 18.9 |
| Immune system clears HPV | True | 377 | **53.9** |
|  | False | 322 | 46.1 |
| HPV infects males | True | 592 | **84.7** |
|  | False | 107 | 15.3 |
| HPV can be asymptomatic | True | 421 | **60.2** |
|  | False | 278 | 39.8 |
| No cure for HPV | True | 453 | 64.8 |
|  | False | 246 | 35.2 |
| HPV vaccine exists | True | 589 | **84.3** |
|  | False | 110 | 15.7 |
| Vaccine protects against multiple types | True | 561 | **80.3** |
|  | False | 138 | 19.7 |
| Both genders should be vaccinated | True | 597 | **85.4** |
|  | False | 102 | 14.6 |
| Vaccine clears existing infection | True | 326 | 46.6 |
|  | False | 373 | **53.4** |
| Screening required before vaccination | True | 528 | **75.5** |
|  | False | 171 | 24.5 |
| Screening required after vaccination | True | 592 | **84.7** |
|  | False | 107 | 15.3 |
| Vaccine recommended for ages 9–14 years | True | 575 | **82.3** |
|  | False | 124 | 17.7 |
| Vaccine costs GHS 1,000–2,500 | True | 403 | 57.7 |
|  | False | 296 | 42.3 |
| Two or more doses required | True | 506 | **72.4** |
|  | False | 193 | 27.6 |
| Condoms protect against HPV | True | 462 | **66.1** |
|  | False | 237 | 33.9 |
| WHO goal: 90% vaccinated by age 15 | True | 597 | **85.4** |
|  | False | 102 | 14.6 |

Note; Data are presented as absolute frequencies and percentages. Abbreviations: HPV: Human papillomavirus.

Regarding the clinical profile and transmission of HPV, respondents demonstrated strong foundational knowledge. A large majority correctly identified the association between HPV and cervical cancer at 87.1% (n=609), recognized its status as a sexually transmitted infection at 88.6% (n=619), and understood that it could infect males at 84.7% (n=592) and cause oropharyngeal cancer at 74.0% (n=517). Furthermore, 69.2% (n=484) of participants correctly rejected the misconception that HPV transmission occurs only in the presence of visible symptoms.

Knowledge surrounding HPV vaccination was similarly robust. Most participants understood that the vaccine is recommended for both genders at 85.4% (n=597), is appropriate for individuals aged 9–14 years at 82.3% (n=575), and provides protection against multiple HPV strains at 80.3% (n=561). Furthermore, 84.7% (n=592) correctly recognized that cervical cancer screening remains necessary even after vaccination.

Despite these high levels of baseline knowledge, critical misconceptions persist. Notably, more than half of the participants 53.4% (n=373) incorrectly believed that the HPV vaccine is effective in treating existing HPV infections, reflecting a significant misunderstanding of the vaccine’s prophylactic nature. Additionally, knowledge regarding preventative measures was suboptimal, with only 66.1% (n=462) of respondents correctly identifying that condom use provides some protection against HPV infection.

### HPV Knowledge Assessment

The combined HPV knowledge score was computed from 21 items assessing participants’ understanding of HPV infection, cervical cancer, and HPV vaccination. Each correct response was awarded one point, while incorrect responses were assigned zero points, resulting in a theoretical score range of 0 to 21. In this study, the observed scores ranged from 3 to 21. Consistent with established literature in this field, a score of 75% or higher was categorized as “good knowledge”(22,23). As shown in Figure 1., the majority of the cohort achieved good knowledge levels, accounting for 51.9% (n=363) of the sample, while 48.1% (n=336) were classified as having poor knowledge.

**Figure 1:**
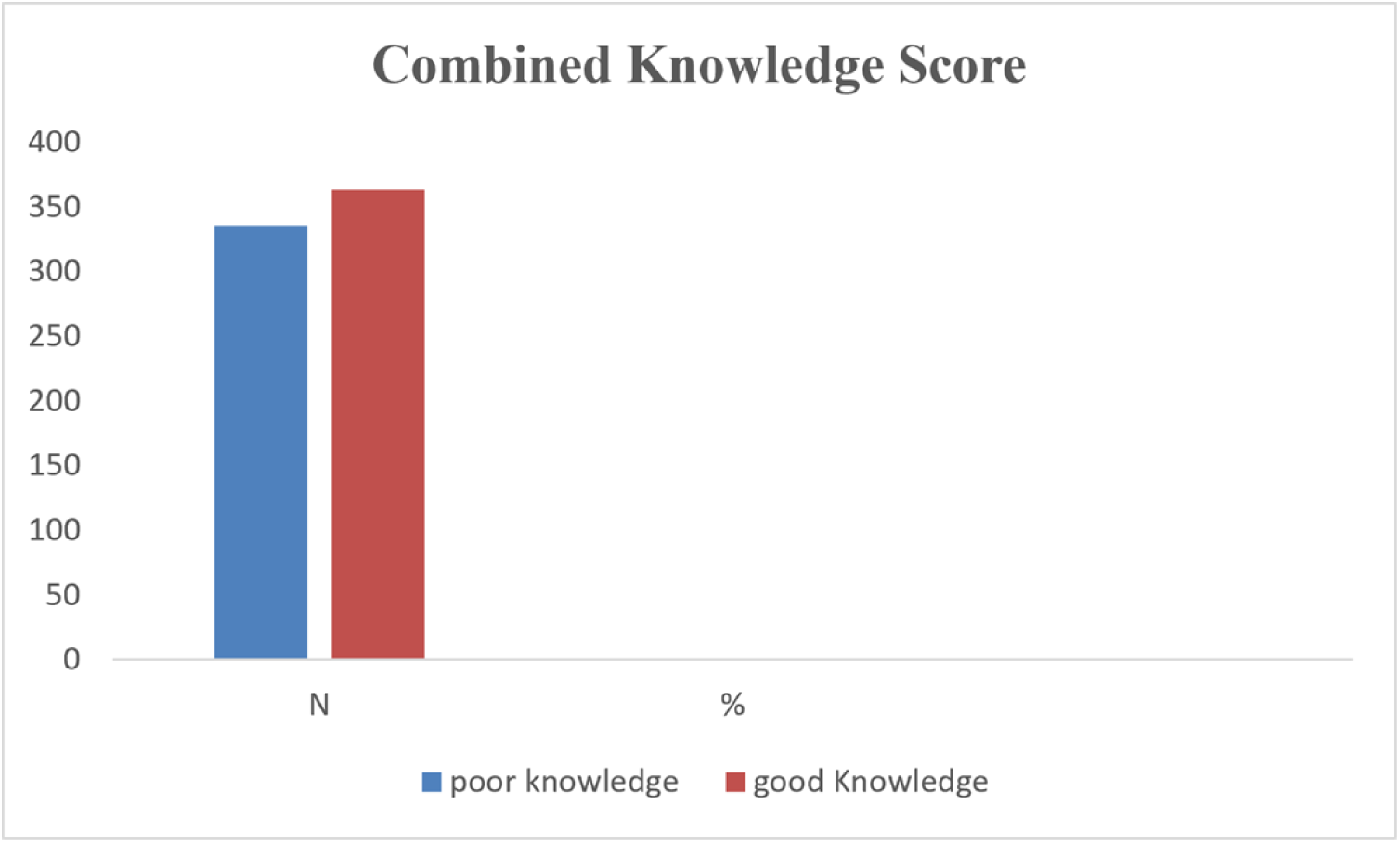
**Combined Knowledge Score of HPV infection, cervical cancer, and HPV vaccination**

### Factors Associated with HPV Knowledge

Bivariate analysis was conducted to identify sociodemographic and behavioral factors associated with HPV knowledge among participants **(Table 3)**. Statistical significance was determined using the Chi-square (X^2^) test, with effect sizes reported using the Cramer’s V coefficient.

**Table 3:** Bivariate Analysis of Factors Associated with HPV Knowledge (N = 699)

| Variable | Poor Knowledge n (%) | Good Knowledge n (%) | $\chi^2$ (df) | p-value | V |
| --- | --- | --- | --- | --- | --- |
| Marital Status | 290 (47.6) | 319 (52.4) | 0.38<br>(1) | .536 | .023 |
| Gender | 173 (45.8) | 205 (54.2) | 1.75<br>(1) | .186 | .050 |
| College Enrolled | 99 (55.6) | 79 (44.4) | 7.86<br>(3) | **.049\*** | .106 |
| Academic Level | 49 (43.0) | 65 (57.0) | 4.91<br>(5) | .427 | .084 |
| Smoking Status | 5 (71.4) | 2 (28.6) | 1.55<br>(1) | .214 | .047 |
| Alcohol Status | 23 (46.9) | 26 (53.1) | 0.03<br>(1) | .870 | .006 |
| Sexual Intercourse | 88 (46.8) | 100 (53.2) | 0.16<br>(1) | .686 | .015 |
| Sexual Partners | 259 (47.6) | 285 (52.4) | 2.39<br>(4) | .665 | .058 |
| Contraceptive Use | 74 (47.4) | 82 (52.6) | 0.03<br>(1) | .858 | .007 |
| Heard of HPV | 241 (44.5) | 301 (55.5) | 12.55<br>(1) | **<.001\*\*\*** | .134 |
| Heard of Cervical Cancer | 312 (47.6) | 344 (52.4) | 1.10<br>(1) | .294 | .040 |
| Heard of Genital Warts | 265 (45.1) | 322 (54.9) | 12.55<br>(1) | **<.001\*\*\*** | .134 |
| Heard of HPV Vaccine | 219 (43.1) | 289 (56.9) | 18.31<br>(1) | **<.001\*\*\*** | .162 |
| Aware of WHO Recommendation | 116 (41.0) | 167 (59.0) | 9.55<br>(1) | **.002\*\*** | .117 |

*Note.* $X^2$ = Pearson chi-square statistic; df = degrees of freedom; V = Cramer's V. \* $p < 0.10$ . \*\* $p < 0.05$ . \*\*\* $p < 0.001$

The analysis revealed that college enrolled in was a significant determinant of HPV knowledge (X^2^ (3) = 7.86, p =.049). Conversely, no significant associations were observed between HPV knowledge and marital status, gender, academic level, smoking status, alcohol consumption, sexual experience, number of sexual partners, or contraceptive use (p >.05).

Participants’ prior exposure to information served as a robust predictor of their current knowledge level. Specifically, those who had previously heard of HPV (X^2^ (1) = 12.55, p <.001), genital warts (X^2^ (1) = 12.55, p <.001), or the HPV vaccine (X^2^ (1) = 18.31, p <.001) were significantly associated with good knowledge. Furthermore, awareness of WHO vaccination recommendations was positively associated with good knowledge scores (X^2^ (1) = 9.55, p =.002).

### Knowledge Levels Regarding HPV and Cervical Cancer by College of Enrollment

The distribution of knowledge levels varied significantly across academic colleges, as illustrated in **Figure 2**. Students in the College of Agriculture and Natural Sciences reported the highest knowledge score (58.8%), followed closely by those in the College of Health and Allied Sciences (54.5%). Conversely, students in the Humanities and Legal Studies college exhibited the lowest proportion of good knowledge (44.4%). These descriptive findings support the bivariate analysis, which confirmed that college enrollment is a significant factor in determining students’ awareness of HPV and cervical cancer, (X^2^ (3) = 7.86, p =.049).

**Figure 2.**
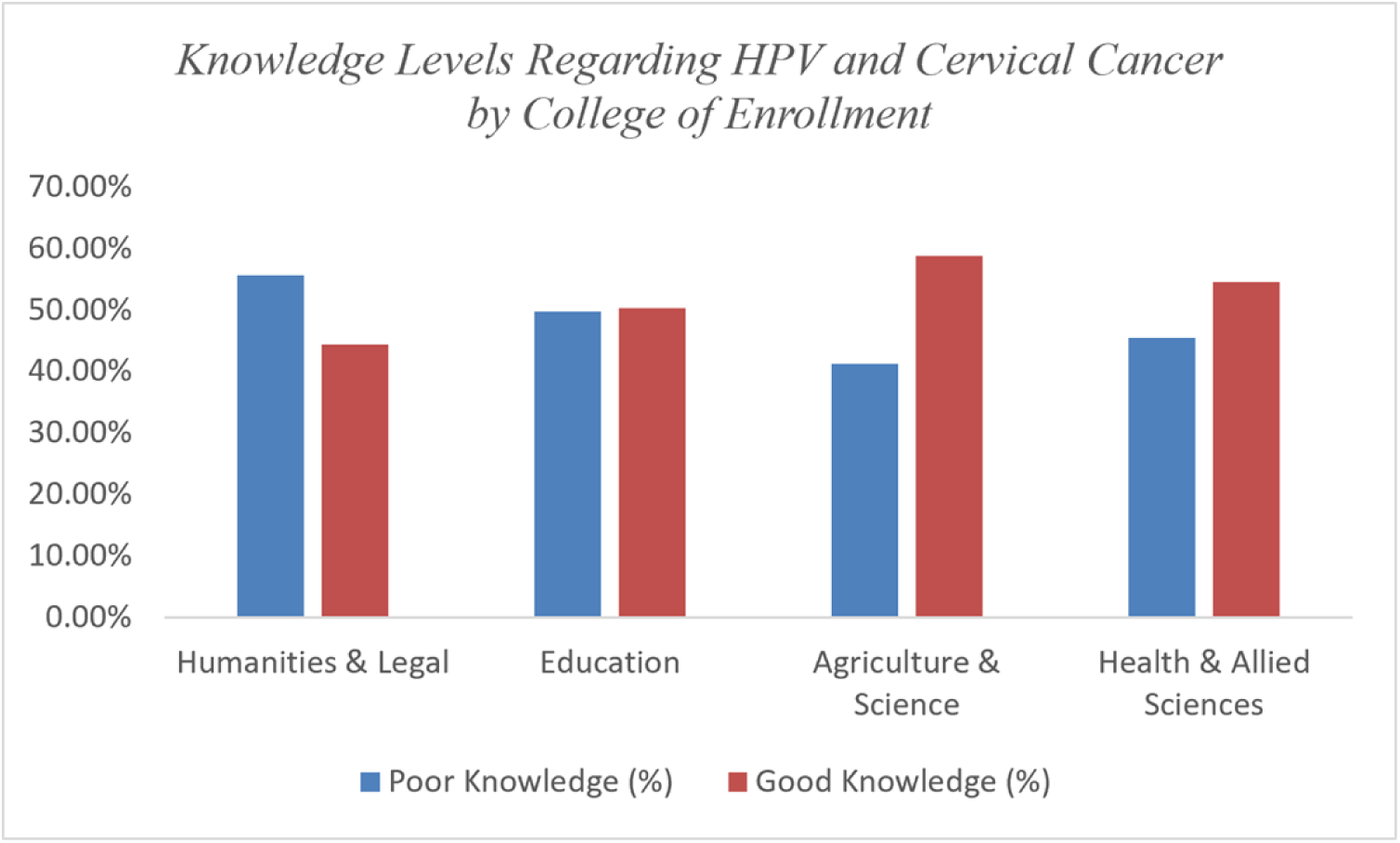
**Distribution of Knowledge Levels Regarding HPV and Cervical Cancer by College of Enrollment**

### Predictors of Good HPV and Cervical Cancer Knowledge

Binary logistic regression was employed to identify independent predictors of good knowledge regarding HPV and cervical cancer **(Table 4).** The model identified three significant independent predictors. Participants who were male were significantly more likely to demonstrate good knowledge compared to their female counterparts (OR = 1.43; 95% CI: 1.04, 1.97; *p* =.026). Furthermore, prior awareness of the HPV vaccine (OR = 1.69; 95% CI: 1.04, 2.75; *p* =.034) and awareness of genital warts (OR = 1.70; 95% CI: 1.02, 2.83; *p* =.043) remained significant predictors of good knowledge when controlling for other covariates. Other variables, including prior awareness of HPV, cervical cancer, WHO recommendations, academic college, and academic level, did not reach statistical significance in the multivariate model.

**Table 4:** Binary Logistic Regression Predicting Good HPV and Cervical Cancer Knowledge (N = 699)

| Predictor | B | SE | OR | 95% CI | p |
| --- | --- | --- | --- | --- | --- |
| **Gender** (Male vs. Female) | 0.36 | 0.16 | 1.43 | [1.04, 1.97] | **.026\*** |
| **Heard of HPV Vaccine** (Yes vs. No) | 0.53 | 0.25 | 1.69 | [1.04, 2.75] | **.034\*** |
| **Heard of Genital Warts** (Yes vs. No) | 0.53 | 0.26 | 1.70 | [1.02, 2.83] | **.043\*** |
| **Heard of HPV** (Yes vs. No) | 0.19 | 0.26 | 1.21 | [0.73, 2.00] | .455 |
| **Heard of Cervical Cancer** (Yes vs. No) | -0.55 | 0.39 | 0.58 | [0.27, 1.23] | .154 |
| **Aware of WHO Recommendation** (Yes vs. No) | 0.24 | 0.18 | 1.27 | [0.90, 1.80] | .168 |
| **College Enrolled** | - | - | - | - | .060 |
| **Academic Level** | - | - | - | - | .378 |

*Note.* B = unstandardized regression coefficient; SE = standard error; OR = odds ratio; CI = confidence interval \* $p < 0.05$ . \*\* $p < 0.01$ . \*\*\* $p < 0.001$ .

### Attitudes Towards HPV, Cervical cancer and HPV Vaccination

Participants’ attitudes toward HPV, cervical cancer and HPV vaccination are summarized in **Table 5**. To assess these attitudes, a composite score was calculated (range: 25–63), with a median score of 41.The median was utilized as the threshold to dichotomize attitudes; scores above 41 were classified as a positive attitude, while scores of 41 or below were classified as a negative attitude (22,24). Based on this categorization, 52.2% (n=365) of participants demonstrated a positive attitude, while 47.8% (n=334) held a negative attitude as shown in **figure 3**.

**Figure 3:**
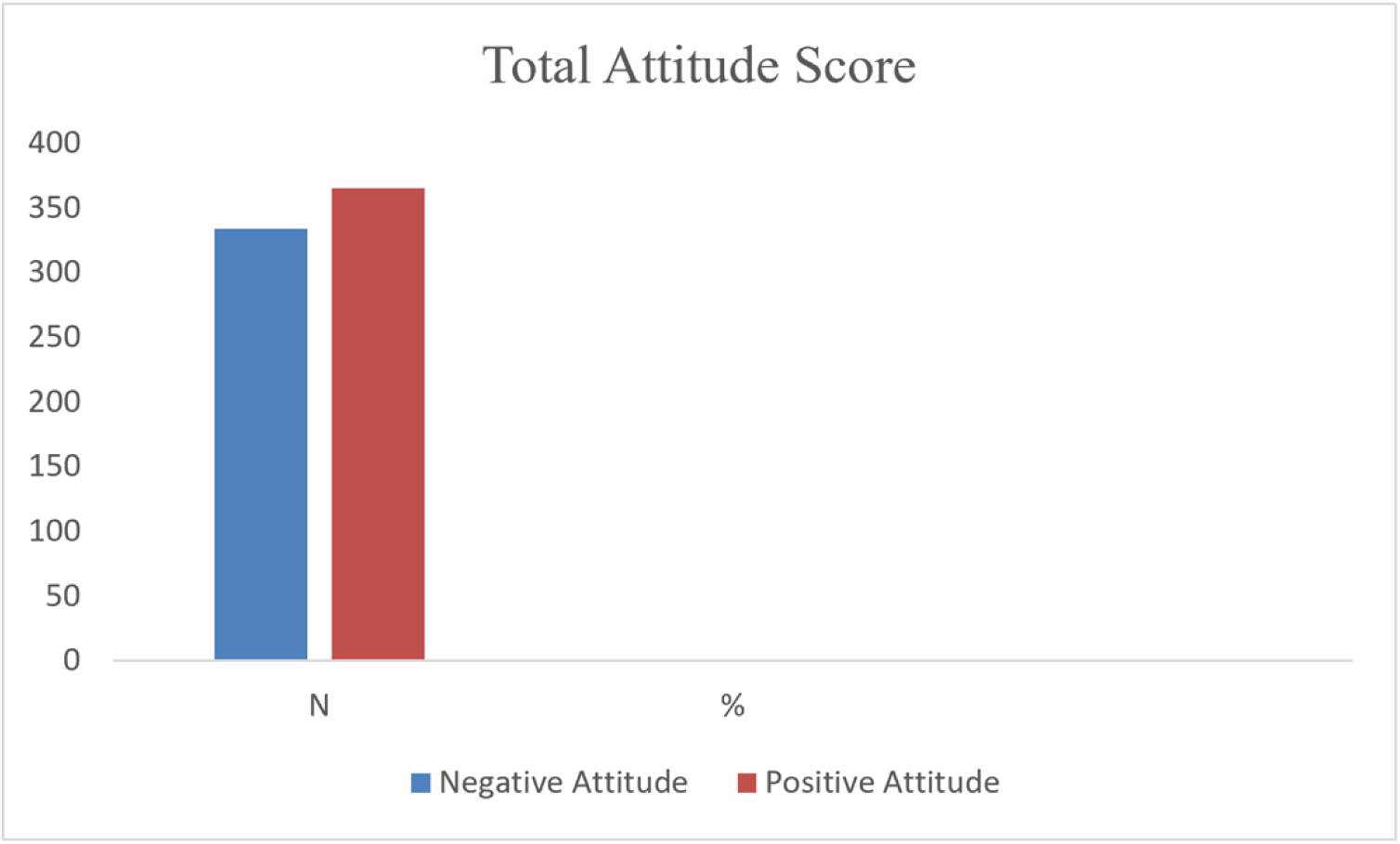
**Total Attitude Score towards HPV and HPV vaccine**

**Table 5:** Participants’ Attitudes Towards HPV Vaccination (n = 699)

| **Attitude Assessing Statement** | **Strongly Agree**<br>**n (%)** | **Agree**<br>**n (%)** | **Neutral**<br>**n (%)** | **Disagree**<br>**n (%)** | **Strongly Disagree**<br>**n (%)** |
| --- | --- | --- | --- | --- | --- |
| I think I can be easily infected by HPV | 115 (16.5) | 174 (24.9) | 185 (26.5) | 112 (16.0) | 113 (16.2) |
| I think HPV infection is a serious disease | 301 (43.1) | 288 (41.2) | 88 (12.6) | 10 (1.4) | 12 (1.7) |
| Getting the HPV vaccine would be beneficial to a teenage girl or boy's future health | 319 (45.6) | 241 (34.5) | 102 (14.6) | 20 (2.9) | 17 (2.4) |
| Getting the HPV vaccine will send a message to teenagers that it is OK to have sex | 102 (14.6) | 175 (25.0) | 191 (27.3) | 139 (19.9) | 92 (13.2) |
| Education on HPV should be implemented at school | 368 (52.6) | 225 (32.2) | 84 (12.0) | 16 (2.3) | 6 (0.9) |
| Men can get involved to prevent their partner from getting cervical cancer | 320 (45.8) | 253 (36.2) | 95 (13.6) | 24 (3.4) | 7 (1.0) |
| People who have only one sexual partner have a low risk of becoming infected with HPV | 202 (28.9) | 273 (39.1) | 154 (22.0) | 49 (7.0) | 21 (3.0) |
| Using a condom can provide 100% protection against HPV infection | 93 (13.3) | 186 (26.6) | 196 (28.0) | 138 (19.7) | 86 (12.3) |
| HPV vaccination is not necessary because a Pap test can be done to rule out cervical cancer | 75 (10.7) | 147 (21.0) | 202 (28.9) | 155 (22.2) | 120 (17.2) |
| It is preferable to vaccinate both men and women against HPV | 244 (34.9) | 269 (38.5) | 141 (20.2) | 23 (3.3) | 22 (3.1) |
| I believe that Government of Ghana must make the HPV vaccine affordable to all adolescents and young adults | 92 (13.2) | 158 (22.6) | 215 (30.8) | 136 (19.5) | 98 (14.0) |
| I have had family/friends sought my opinion regarding HPV vaccination | 77 (11.0) | 142 (20.3) | 198 (28.3) | 131 (18.7) | 151 (21.6) |
| I believe that the HPV vaccine should be recommended for all teenage girls and young women | 96 (13.7) | 156 (22.3) | 190 (27.2) | 121 (17.3) | 136 (19.5) |
| I believe that vaccination should be recommended for all teenage boys and young men | 113 (16.2) | 177 (25.3) | 203 (29.0) | 105 (15.0) | 101 (14.4) |
| I believe it would be difficult and embarrassing for me to ask for the HPV vaccine because it is associated with a sexually transmitted infection | 187 (26.8) | 226 (32.3) | 208 (29.8) | 50 (7.2) | 28 (4.0) |
| If other people knew I received the HPV vaccine, I would be embarrassed | 199 (28.5) | 259 (37.1) | 183 (26.2) | 40 (5.7) | 18 (2.6) |
| I would worry about what my partner or future partner would think if I received the HPV vaccine | 298 (42.6) | 247 (35.3) | 118 (16.9) | 25 (3.6) | 11 (1.6) |

Regarding specific attitudinal statements, a strong recognition of disease severity was evident, with 84.3% (n=589) of participants agreeing or strongly agreeing that HPV infection is a serious disease. However, perceptions of personal susceptibility were lower, as only 41.4% (n=289) believed they were personally at risk of infection. Support for institutional health education was robust, with 84.8% (n=593) of respondents favoring the implementation of HPV education in schools.

Despite this baseline support, participants reported significant psychosocial barriers. Stigma emerged as a primary concern: 59.1% (n=413) of participants agreed or strongly agreed that requesting the vaccine would be embarrassing due to its association with an STI. Furthermore, 65.6% (n=458) indicated they would feel embarrassed if their vaccination status were known, and 77.9% (n=545) expressed apprehension regarding how a current or future partner might perceive them if they were vaccinated.

Finally, the study identified persistent misconceptions that may hinder preventive behaviors. Approximately 39.9% (n=279) of participants incorrectly believed that condom use provides 100% protection against HPV, and 68.0% (n=475) erroneously believed that having only one sexual partner significantly mitigates the risk of infection. Additionally, 31.7% (n=222) incorrectly perceived the HPV vaccine as unnecessary for individuals undergoing routine cervical cancer screening.

### Factors Associated with Attitudes Toward HPV and Cervical Cancer

Bivariate analysis was conducted to identify sociodemographic and behavioral correlates of participants’ attitudes toward HPV and cervical cancer **(Table 6).** Significant associations were identified between gender and attitudes, with female students being significantly more likely to express a positive attitude at 62.3% (n=200) compared to male students at 43.7% (n=165) (X^2^ (1) = 24.21, p <.001). Furthermore, prior health information exposure was significantly associated with positive attitudes. Participants who had previously heard of HPV were significantly more likely to have positive attitudes at 55.4% (n=300) compared to those who had not (41.4%, n=65) (X^2^ (1) = 9.49, p =.002). Similarly, positive attitudes were significantly more prevalent among those who had heard of genital warts (54.7%, n=321 X^2^ (1) = 8.94, p =.003), the HPV vaccine (55.1%, n=280; X^2^ (1) = 6.27, p =.012), and cervical cancer (53.2%, n=349; X^2^ (1) = 4.14, p =.042). No significant associations were found between attitudes and other variables, including marital status, college enrollment, academic level, substance use, or sexual history (p >.05).

**Table 6:**
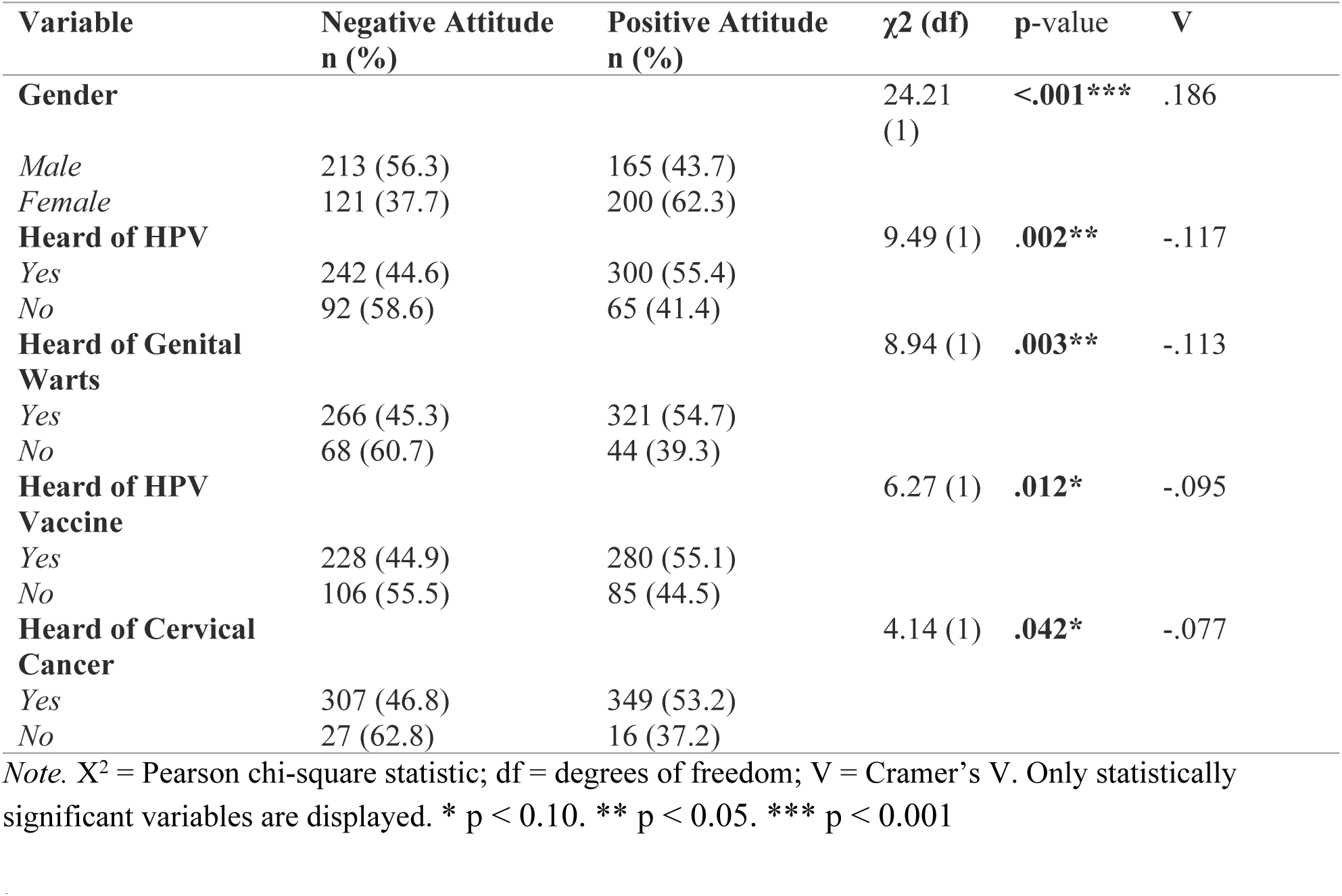
Bivariate Analysis of Factors Associated with Attitude Toward HPV and Cervical Cancer (N = 699)

### Attitudes Toward HPV and Cervical Cancer between Male And Female

As illustrated in Figure 4, attitudes toward HPV and cervical cancer differed significantly by gender (X^2^ (1) = 24.21, p <.001). Most female students reported a positive attitude at 62.3% (n=200), whereas male students were more likely to express a negative attitude at 56.3% (n=213). These findings indicate that gender is a significant determinant of student perspectives regarding HPV and cervical cancer awareness, suggesting that while male students may possess higher baseline knowledge, female students demonstrate a higher degree of attitudinal favorability toward HPV vaccination.

**Figure 4:**
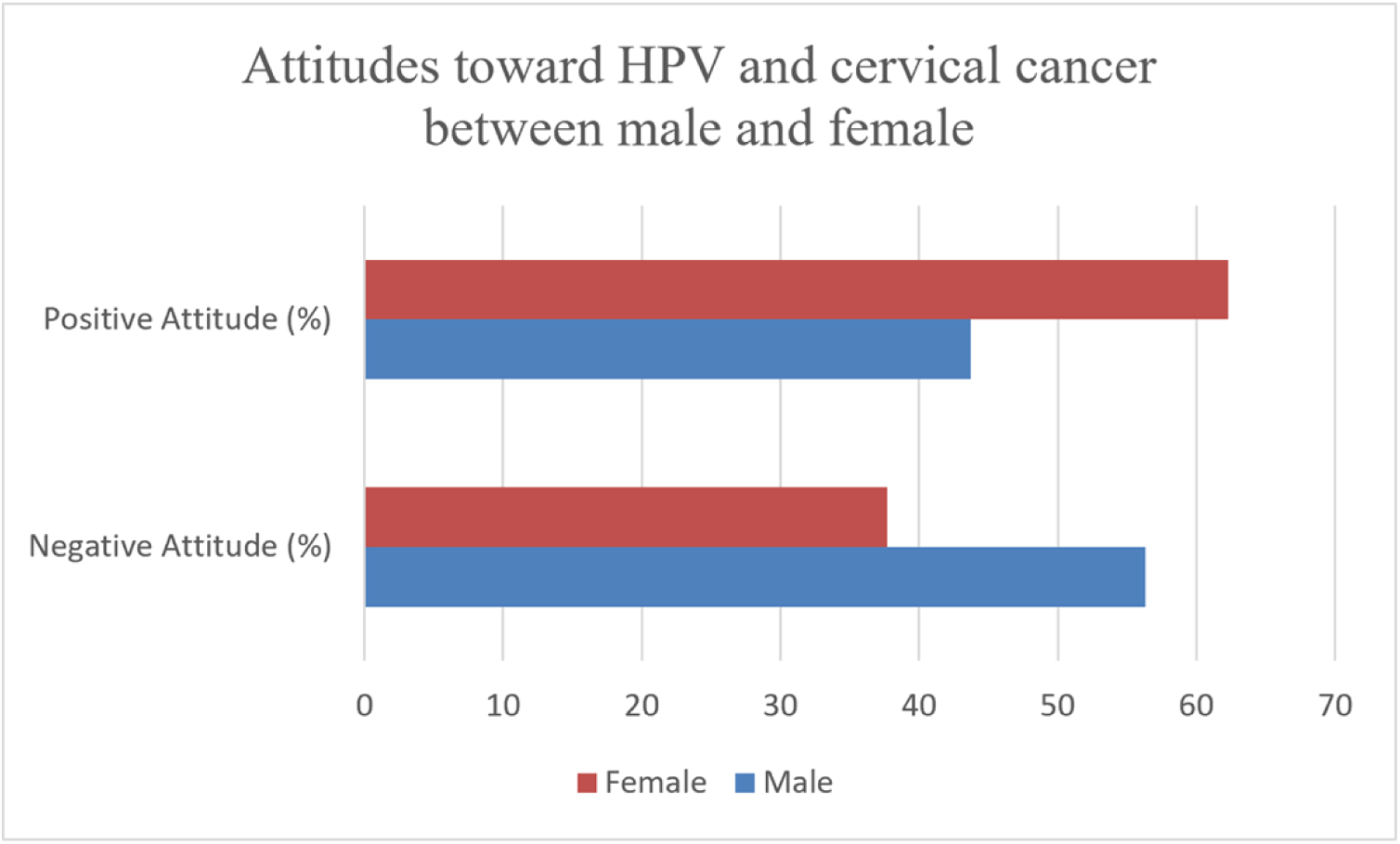
**Comparison of attitudes toward HPV and cervical cancer between male and female undergraduate students.**

### Predictors of Positive Attitudes Toward HPV and Cervical Cancer

Binary logistic regression analysis was conducted to determine the independent predictors of a positive attitude toward HPV and cervical cancer prevention **(Table 7).** The analysis revealed that gender was the only significant independent predictor of attitude. Male students were 50% less likely to report a positive attitude toward HPV and cervical cancer prevention compared to female students (OR = 0.50; 95% CI: 0.36, 0.69; *p* <.001). Other variables included in the model including prior awareness of HPV, genital warts, the HPV vaccine, and WHO recommendations did not reach statistical significance after controlling for other factors (p >.05).

**Table 7:**
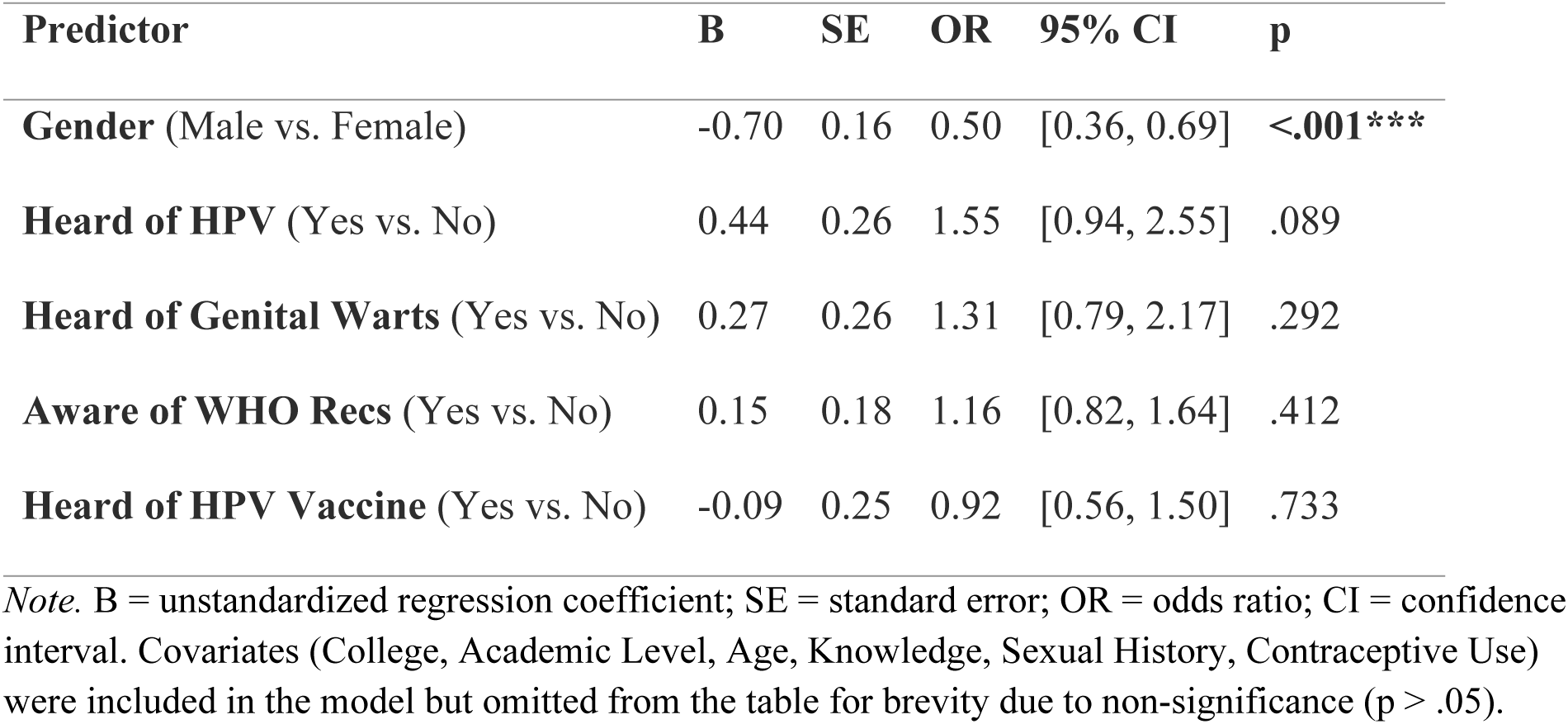
Binary Logistic Regression Predicting Positive Attitude Toward HPV and Cervical Cancer (N = 699)

| Predictor | B | SE | OR | 95% CI | p |
| --- | --- | --- | --- | --- | --- |
| Gender (Male vs. Female) | -0.70 | 0.16 | 0.50 | [0.36, 0.69] | <.001*** |
| Heard of HPV (Yes vs. No) | 0.44 | 0.26 | 1.55 | [0.94, 2.55] | .089 |
| Heard of Genital Warts (Yes vs. No) | 0.27 | 0.26 | 1.31 | [0.79, 2.17] | .292 |
| Aware of WHO Recs (Yes vs. No) | 0.15 | 0.18 | 1.16 | [0.82, 1.64] | .412 |
| Heard of HPV Vaccine (Yes vs. No) | -0.09 | 0.25 | 0.92 | [0.56, 1.50] | .733 |

*Note.* B = unstandardized regression coefficient; SE = standard error; OR = odds ratio; CI = confidence interval. Covariates (College, Academic Level, Age, Knowledge, Sexual History, Contraceptive Use) were included in the model but omitted from the table for brevity due to non-significance ( $p > .05$ ).

### Perceived Barriers to HPV Vaccination

The barriers to HPV vaccination identified among the study population are illustrated in **Figure 5** These barriers were categorized into Cognitive gaps, psychosocial/stigma, perceived susceptibility and gender-based disparities.

**Figure 5:**
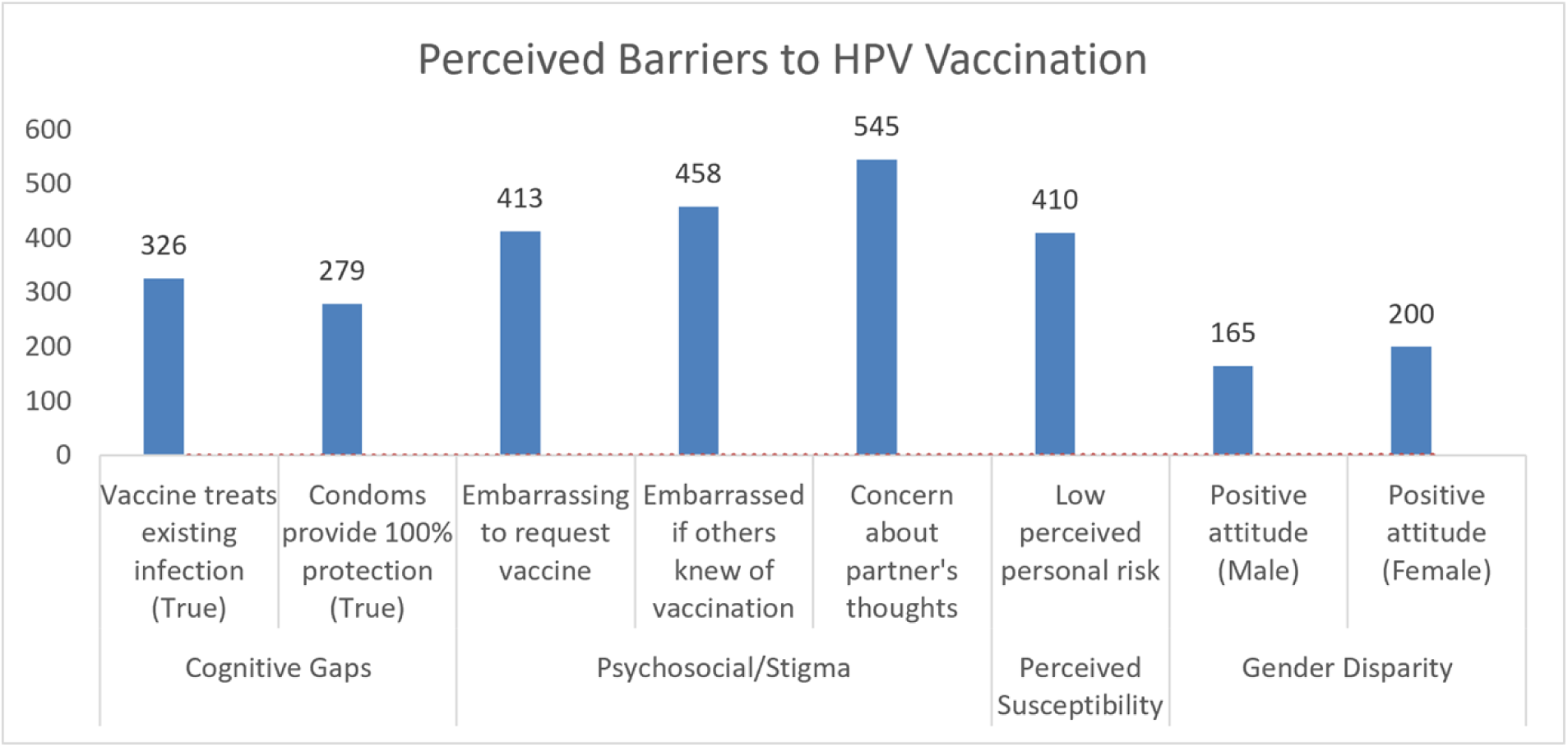
**Perceived Barriers to HPV Vaccination**

Cognitive barriers were prevalent, with 46.6% (n=326) of participants incorrectly believing that the HPV vaccine treats existing infections, while 39.9% (n=279) mistakenly identified condom use as providing 100% protection against HPV. Psychosocial concerns represented the most significant obstacles; a substantial majority of 77.9% (n=545) expressed apprehension regarding their partner’s perception if they were vaccinated. Furthermore, stigma surrounding the sexual nature of the virus was a major deterrent, with 65.6% (n=458) reporting embarrassment if others discovered they had received the vaccine, and 59.1% (n=413) noting that requesting the vaccine would be difficult due to its association with STIs. Finally, low perceived susceptibility was a significant behavioral barrier, with 58.6% (n=410) of participants reporting that they did not feel they were personally at risk for HPV infection.

## Discussion

This study assessed the knowledge, attitudes, and barriers regarding HPV, cervical cancer and HPV vaccination among undergraduate students at the University of Cape Coast. While general awareness of these conditions is present, our findings underscore a paradoxical disconnect high recognition of disease severity is simultaneously hampered by profound psychosocial stigma and fundamental cognitive misconceptions. This study offers a comprehensive look at how individual, gender and systemic factors shape preventive health perspectives.

### The Knowledge-Attitude Paradox and Gender Perspectives

Our findings revealed that 51.9% of the participants possessed good knowledge, a figure higher than that reported among tertiary students in Nigeria where good knowledge generally ranged from about 5–32% (25,26). Awareness of HPV (77.5%) and cervical cancer (93.8%) was higher than in several university-based studies from Turkey, Pakistan, Saudi Arabia, and Ethiopia, where a substantial proportion of students had low of awareness of HPV or the vaccine (27–30). Additionally, studies conducted among Ghanaian women reported low knowledge levels (31,32), for instance Nartey et al. (2024) reported that fewer than 10% of women were aware of HPV infection, and only 4% knew about HPV vaccination. A study conducted at the University of the Free State in Bloemfontein, South Africa, among female students reported that out of 381 participants, only 40.9% of participants had good knowledge of HPV infection, only 9.7% demonstrated good knowledge of HPV vaccination, and 13.3% had good overall knowledge (13). These results suggest that, baseline exposure to HPV-related information is higher among undergraduates in the University of Cape Coast. This may be attributable to formal education, access to health information via the University Hospital, and exposure to recent national vaccination campaigns, including the nationwide HPV vaccination initiative launched in 2025 (4)

However, a striking gender divergence emerged. While our logistic regression analysis indicated that male students were 1.43 times more likely to demonstrate good knowledge than their female counterparts, female students were significantly more likely to hold a positive attitude toward vaccination (p <.001). This findings contrast contrasts with studies from Turkey, Saudi Arabia, Italy, and multiple LMIC settings where women generally show higher HPV knowledge (29,33–35).

This discrepancy suggests a knowledge-attitude paradox. Male students may perceive HPV knowledge as a clinical or academic fact potentially reinforced by biology-focused curricula whereas female students, recognizing the direct risk of cervical cancer, likely frame this information through a lens of personal health relevance. Our findings generate a new hypothesis for the Ghanaian context: that health literacy alone is insufficient for men, as they may lack the perceived personal susceptibility necessary to transform clinical knowledge into a positive attitudinal stance.

### Psychosocial Stigma as a Primary Barrier

The most compelling finding of this study is the influence of barriers, which persist despite participants’ high acknowledgement of HPV’s severity (84.3%). Nearly 78% of students expressed apprehension regarding partner perceptions, and 65.6% reported fear of embarrassment if their vaccination status were disclosed. Similar concerns have been documented among adolescents and young adults in Nigeria, where HPV vaccination is sometimes perceived as an indicator of sexual activity or promiscuity (36). This aligns with the Health Belief Model (HBM), where perceived social barriers are often stronger predictors of behavioral intent than the actual threat of disease(37).

Unlike in many Western contexts where the barrier to HPV vaccination is often centered on vaccine hesitancy(14,38,39), our data suggests that for Ghanaian undergraduates, the barrier is fundamentally rooted in the stigma associated with STIs. This has critical implications for clinical practice: standard clinical delivery models which prioritize medical efficacy are inadequate if the delivery environment is perceived as socially compromising. We propose that to improve uptake, vaccination strategies must be integrated into general campus health services rather than specialized reproductive clinics, thereby masking the STI-specific stigma.

### Cognitive Misconceptions and Educational Implications

The presence of persistent misconceptions such as the belief that condoms provide 100% protection or that the vaccine treats existing infections suggests that current information sources, often limited to social media, lack the nuance of clinical education. As our regression analysis confirmed, prior awareness is insufficient for positive behavioral outcomes; structured education is required. Consistent with evidence from similar demographics in Bangladesh and India, the lack of standardized HPV-related curriculum in universities leaves a vacuum filled by misinformation(20,40). Additionally, Similar misunderstandings have been reported among university populations in Nigeria and Ethiopia, where overreliance on screening and misconceptions regarding sexual risk contributed to lower vaccine uptake(41,42).

Ultimately, by identifying the specific psychosocial drivers of vaccine hesitancy among university students, our findings provides a localized evidence essential for refining campus-based immunization strategies, thereby contributing to the broader World Health Organization 2030 goal of eliminating cervical cancer through high-coverage, stigma-neutral vaccination programs.

### Implications for Practice, Research, and Policy

Clinical practice must shift toward a stigma-neutral model of care to overcome the identified psychosocial barriers. Currently, the delivery of the HPV vaccine is often siloed within reproductive health or STI clinics, which inadvertently reinforces the social embarrassment reported by over 60% of our participants. Clinicians should integrate HPV immunization into routine primary care or general student health screenings, framing the vaccine strictly as a cancer-prevention tool rather than an STI-preventive measure. By normalizing the vaccine alongside other routine immunizations such as those for Hepatitis B or Tetanus healthcare providers can reduce the social anxiety and perceived stigma that currently prevent students from seeking the vaccine. Future research is required to move beyond descriptive cross-sectional designs to determine the long-term drivers of vaccine hesitancy in the Ghanaian context. We recommend the development and evaluation of longitudinal cohort studies that track university students from matriculation to graduation, measuring the transition from knowledge to vaccine uptake while controlling for socio-economic variables. Furthermore, there is a clear need for randomized controlled trials (RCTs) that test the effectiveness of gender-inclusive, peer-led educational interventions. These studies should employ Structural Equation Modeling (SEM) to disentangle the indirect pathways between knowledge, social stigma, and behavioral intention, providing a roadmap for tailoring health communication strategies to specific demographic subgroups.

Policy efforts must prioritize the institutionalization of comprehensive sexual and reproductive health education within the national academic curriculum to address deep-seated cognitive misconceptions. Current awareness campaigns remain insufficient because they lack the depth to correct fundamental errors regarding vaccine efficacy and the nature of HPV infection. National health policy should mandate that university health services receive dedicated funding to provide subsidized, confidential, and accessible HPV vaccination programs. By embedding HPV education into the university experience and removing financial and access-related barriers, policymakers can transform HPV vaccination from an optional, stigma-prone decision into a standard, accessible component of the national strategy for cervical cancer elimination.

### Strengths, Limitations, and Future Research

This study possesses several key strengths. First, we included both male and female participants, addressing a critical gap in existing literature, which has traditionally focused exclusively on the female population. Given that male vaccination and support are essential for broader community immunity and the reduction of HPV transmission, this inclusive approach provides a more comprehensive, gender-balanced perspective on the psychosocial barriers to vaccine uptake. Additionally, this study’s primary strength is its robust, multidisciplinary large sample size (N=699) and the use of multistage stratified random sampling which provided a high level of statistical power to assess associations through both bivariate and multivariate analyses.

However, we acknowledge limitations. The cross-sectional design restricts our ability to infer causal relationships or track longitudinal changes in vaccination intention. Furthermore, because the study was conducted at a single university, findings may not be fully generalizable to non-academic populations or students in other institutional settings.

## Conclusion

This study demonstrates that undergraduate students at the University of Cape Coast generally possess good knowledge toward HPV infection, cervical cancer and vaccination. The current gold standard for cervical cancer prevention remains primary vaccination. However, our findings suggest that in the Ghanaian undergraduate context, the last mile of vaccine uptake is not blocked by a lack of clinical information, but by social anxiety and gendered perceptions of risk. Clinical and public health interventions must shift from purely pedagogical models to holistic strategies that treat stigma as a medical barrier. By decoupling the HPV vaccine from the narrow label of an STI vaccine, health providers can foster an environment where students feel safe prioritizing their long-term health over immediate social perception.

## Data Availability

The minimal data set and survey instrument underlying the findings of this study are available at Figshare (DOI: 10.6084/m9.figshare.32763999)

## Abbreviations

CI: Confidence Interval
GAVI: Global Alliance for Vaccines and Immunization
HBM: Health Belief Model
HPV: Human Papillomavirus
KAP: Knowledge, Attitude and Practice
LMICs: Low-and Middle-Income Countries
OR: Odds Ratio
SD: Standard Deviation
SPSS: Statistical Package for the Social Sciences
SSA: Sub-Saharan Africa
STI: Sexually Transmitted Infection
UCC: University of Cape Coast
UNICEF: United Nations Children’s Fund
WHO: World Health Organization

## Declarations

### Ethics approval and consent to participate

The study protocol was reviewed and approved by the Committee on Human Research, Publication and Ethics (CHRPE) at the Kwame Nkrumah University of Science and Technology (Ref ID: 266072615210). All procedures were conducted in strict accordance with the ethical principles outlined in the Declaration of Helsinki. Prior to data collection, informed consent was obtained from all participants. Potential participants were fully briefed on the study’s objectives, the voluntary nature of their involvement, and their absolute right to withdraw at any time without consequence.

### Consent for publication

Not applicable. This study does not contain any individual person’s data in any form (including personal details, images, or videos).

## Acknowledgements

The authors express their sincere gratitude to all the students who participated in this study. We also extend our appreciation to Kelvin Asemnyra, Joyous Ocran, Bernard Badengo, and Davida Adwoa Asmah for their invaluable assistance during the data collection process.

## Authors’ contributions

I.A.E conceptualized the study. Methodology was developed by I.A.E, C.A, A.A and C.E.B. A.E.D., B.A., T.K.N and E.K.Q collected the data. Formal analysis was conducted by C.E.B. and I.A.E.

I.A.E wrote the original draft. A.A, M.A.E and I.A.E reviewed and edited the manuscript. All authors have read and agreed to the published version of the manuscript.

